# Inequalities in the decline and recovery of pathological cancer diagnoses during the first six months of the COVID-19 pandemic: a population-based study

**DOI:** 10.1101/2021.02.23.21252276

**Authors:** Ashleigh C. Hamilton, David W. Donnelly, Maurice B. Loughrey, Richard C. Turkington, Colin Fox, Deirdre Fitzpatrick, Ciaran E. O’Neill, Anna T. Gavin, Helen G. Coleman

**Affiliations:** Centre for Public Health, Queen’s University Belfast, Belfast, Northern Ireland; Northern Ireland Cancer Registry, Belfast, Northern Ireland; Patrick G. Johnston Centre for Cancer Research, Queen’s University Belfast, Belfast, Northern Ireland; Department of Pathology, Belfast Health and Social Care Trust, Belfast, Northern Ireland

**Keywords:** Health inequalities, epidemiology, cancer trends, cancer registration

## Abstract

**Background:** The restructuring of healthcare systems to cope with the demands of the COVID-19 pandemic has led to a reduction in clinical services such as cancer screening and diagnostics.

**Methods:** Data from the four Northern Ireland pathology labs was used to assess trends in pathological cancer diagnoses from 1^st^ March to 12^th^ September 2020 overall and by cancer site, gender and age. These trends were compared to the same timeframe from 2017-2019.

**Results:** Between 1^st^ March and 12^th^ September 2020 there was a 23% reduction in cancer diagnoses compared to the same time period in the preceding three years. Although some recovery occurred in August and September 2020, this revealed inequalities across certain patient groups. Pathological diagnoses of lung, prostate and gynaecological malignancies remained well below pre-pandemic levels. Males and younger/middle-aged adults, particularly the 50-59 year old patient group, also lagged behind other population demographic groups in terms of returning to expected numbers of pathological cancer diagnoses.

**Conclusions:** There is a critical need to protect cancer diagnostic services in the ongoing pandemic to facilitate timely investigation of potential cancer cases. Targeted public health campaigns may be needed to reduce emerging inequalities in cancer diagnoses as the COVID-19 pandemic continues.

## Background

The COVID-19 pandemic is an unprecedented global challenge that has had profound effects on healthcare systems. The rapid transformation of clinical services across the world, such as the opening of Nightingale Hospitals in the UK, has required a significant effort from healthcare personnel and governments. The redeployment of staff and resources directed towards managing COVID-19 has had a significant impact on provision of routine medical care, with many such services being reduced or cancelled.

Screening for breast, cervical and colorectal cancer was paused across the UK, and in Northern Ireland this occurred from the second week of March 2020 for 4 months.^(1)^ Furthermore, emergency department attendances also dropped by approximately 30% in March 2020 compared to March 2019,^(2)^ illustrating a reduction in health seeking behaviour by the general public. In addition, a national shift to remote clinics and telephone consultations has reduced face-to-face time between patients and healthcare professionals.^(3)^ A national lockdown was instigated from the 23^rd^ March, accompanied by various levels of public restrictions, and public health messaging about protecting the NHS has been prominent in the UK throughout 2020. All of these factors have implications for cancer care, where early diagnosis and timely treatment are essential in improving outcomes.

Attention is now broadening from the acute concern of COVID-19, to recognition of the collateral damage and excess deaths from other causes that may be a consequence of the necessary changes to healthcare systems as a result of the virus. Of major concern are the delays in cancer diagnoses. Predicted trends of later stage at diagnosis of cancer are likely to lead to poorer outcomes for patients, thus reversing years of progress in improving cancer survival.^(4)^ Two modelling studies have estimated the number of cancer deaths as a result of delays in diagnosis during the COVID-19 pandemic in England. One study investigated breast, lung, colorectal and oesophageal cancer diagnosis via urgent and emergency care pathways, and estimated 3291-3621 additional deaths in England over the next five years from these cancers.^(5)^ Another study specifically assessed the effect of delays in the two week wait referral pathway for the 20 most common types of cancer. It was estimated that 181-542 additional lives would be lost as a result of a delay in presentation.^(6)^ Whilst figures from these studies differ due to variation in methodology and cancer sites studied, the estimated loss of life from cancer resulting from the COVID-19 pandemic based on these models is considerable.

Provisional data from the Netherlands Cancer Registry found the number of cancer cases dropped by 9-27% per week from the 24^th^ February to 12^th^ April 2020 compared to the pre-pandemic period.^(7)^ A subsequent modelling study estimated the effect of suspending colorectal and breast cancer screening programmes in the Netherlands during the initial stages of the pandemic. They found a reduction in observed breast and colorectal cancer diagnoses in the population as a whole, but particularly in the screening age groups, with recovery to expected levels by late June 2020.^(8)^ In contrast to this, a Korean study of lung cancer cases in three hospitals found that the number of lung cancer diagnoses from February to June 2020 did not differ significantly from the previous 3 years.^(9)^ Finally, a Danish study investigating all-cause mortality rates (including cancer mortality) from 1^st^ January through to 5^th^ July 2020 found that there was no excess mortality as a result of COVID-19 or the resulting lockdown in Denmark.^(10)^ This is in contrast to several other European countries, including the UK, which had shown excess all-cause mortality by May 2020.^(11)^

Given the findings of modelling studies^(5, 6)^ and limited data published to date, there remains an urgent need to quantify the impact of COVID-19 on cancer diagnoses using real-world data. The aim of this study was to evaluate the impact of COVID-19 on pathological diagnoses of cancer using data collated by the population-based Northern Ireland Cancer Registry (NICR). A secondary aim was to evaluate potential inequalities in the pathological cancer diagnosis trends observed by subgroups of cancer type, patient age, sex and for screen-detected cancer.

## Methods

The NICR is a population-based register covering approximately 1.8 million people, and has collected information on all patients diagnosed with cancer in Northern Ireland since 1993. The NICR is the officially recognised provider of cancer statistics for Northern Ireland, and the high quality of the NICR data, including its validity and completeness have been previously reported.^(12)^ Ethical approval for the NICR databases (including the waiving of requirement for individual patient consent) and for data analysis has been granted by the Office for Research Ethics Committees of Northern Ireland (ORECNI reference 15/NI/0203), recently renewed in October 2020 (ORECNI reference 20-NI-0132).

Data from the four NHS pathology laboratories in Northern Ireland are usually provided to the NICR on a monthly basis. These data were used to compile a summary of trends in the number of patients with pathology samples indicating cancer, from 1^st^ March to 12^th^ September 2020 during the COVID-19 pandemic. This time period reflects both the temporary suspension of breast, colorectal and cervical screening programmes, which were paused for four months from March 2020, as well as referral pathways for symptomatic patients. Data was recorded using the number of pathology samples indicating cancer. In the instance of an individual having more than one pathology sample, patients were only included once per cancer type unless they have had pathology samples separated by more than two years. The NICR produces monthly reports of these trends, which are available online.^(13)^ The methodology used in this study differs from usual NICR registration practices which collects information electronically from a number of sources including the Pathology Laboratory System alongside the hospital Patient Administration System, multidisciplinary team meeting system, Death Certificates and Radiology Systems.

### Statistical analysis

Descriptive statistics (frequencies and proportions over time) are presented for the number of patients diagnosed with cancer, excluding non-melanoma skin cancer, in Northern Ireland between 1^st^ March and 12^th^ September 2020. Comparisons were made to the same week range for 2017-2019, for which a three year average was estimated. Data during the most recent five weeks for which data was available, from 9^th^ August to 12^th^ September 2020, was also evaluated separately to assess for any signs of recovery in pathological cancer diagnoses. Subgroup analysis was conducted by age, sex and cancer site, including screen-detected cancers. For analysis by age category, we have restricted the data to adults aged 18 years and older. The monthly trend in patients diagnosed pathologically from 2017 to 2019 was used to estimate the number of patients that would normally be expected to have a pathology sample indicating cancer during March-August 2020. An estimate of the number of ‘missed’ patients was then based upon the difference between the observed and expected number of patients recorded over this time period.

## Results

During the time period from 1^st^ March through to 12^th^ September 2020, there were 3,561 pathology samples indicating a cancer diagnosis in people of all ages in Northern Ireland. During the same time period in 2017-2019 there were, in each year, an average number of 4,607 pathology samples indicating a cancer diagnosis. This corresponds to a 22.7% reduction in cancer cases diagnosed during the six month peak period of the first wave of the COVID-19 pandemic in Northern Ireland. Based on monthly trends, there was an estimated absolute shortfall of 1,130 patients during March to August 2020 inclusive, compared to the same period for 2017-2019 (Figure 1). In April 2020, new cancer diagnoses reached their lowest point during the pandemic, with a decrease of 45.2%, but some signs of recovery were evident by June 2020.

**Figure 1.**
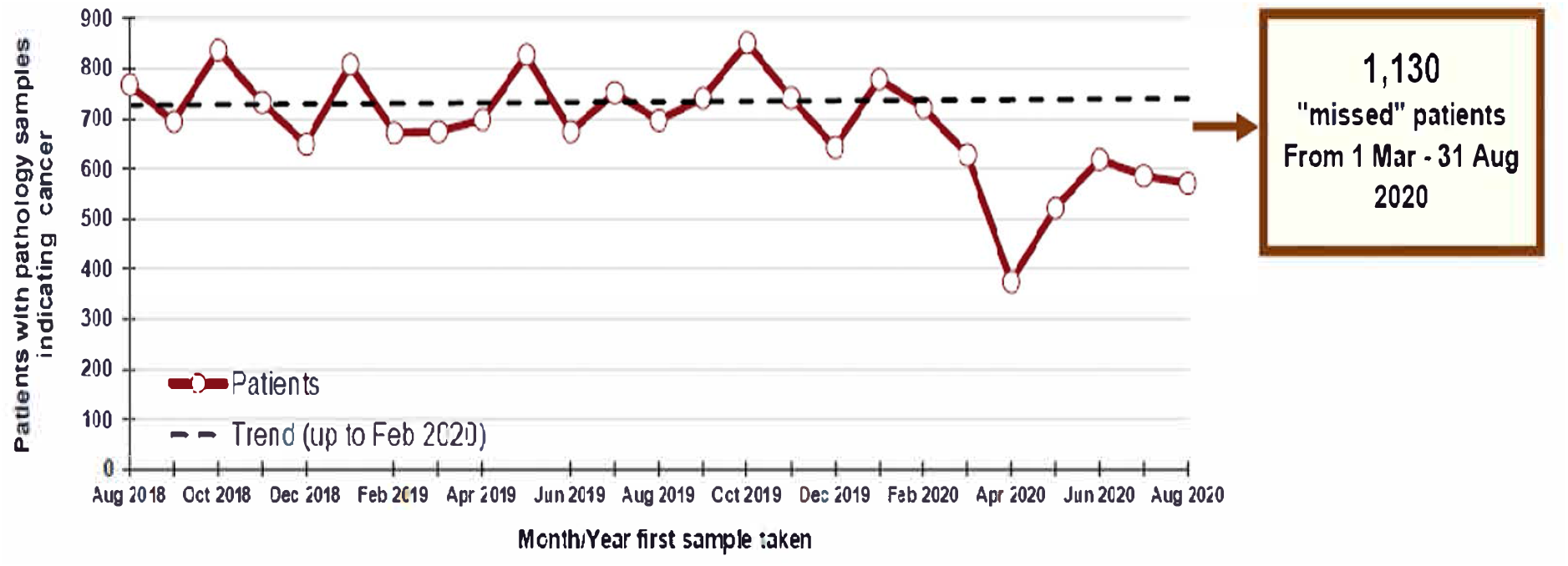
Trend in Patients with Pathology Samples Indicating Cancer by Month and Year First Sample Taken. Reproduced from the Northern Ireland Cancer Registry website^(13)^

### Analysis by cancer site

The distribution of cancer diagnoses by site is shown in Figure 2. The number of diagnoses across all cancer sites was reduced during the overall period from 1^st^ March to 12^th^ September 2020 compared to the same time period in the preceding three years. However, in August and September, there were signs of recovery in the number of diagnoses across several cancer sites, with colorectal cancer, breast cancer and haematological malignancies showing a 12%, 6% and 30% increase respectively compared to previous years. Of concern though, lung, prostate and gynaecological cancer diagnoses remained considerably lower than expected during this most recent five week time frame, at 21%, 10% and 18% respectively below expected numbers of pathological diagnoses.

**Figure 2.**
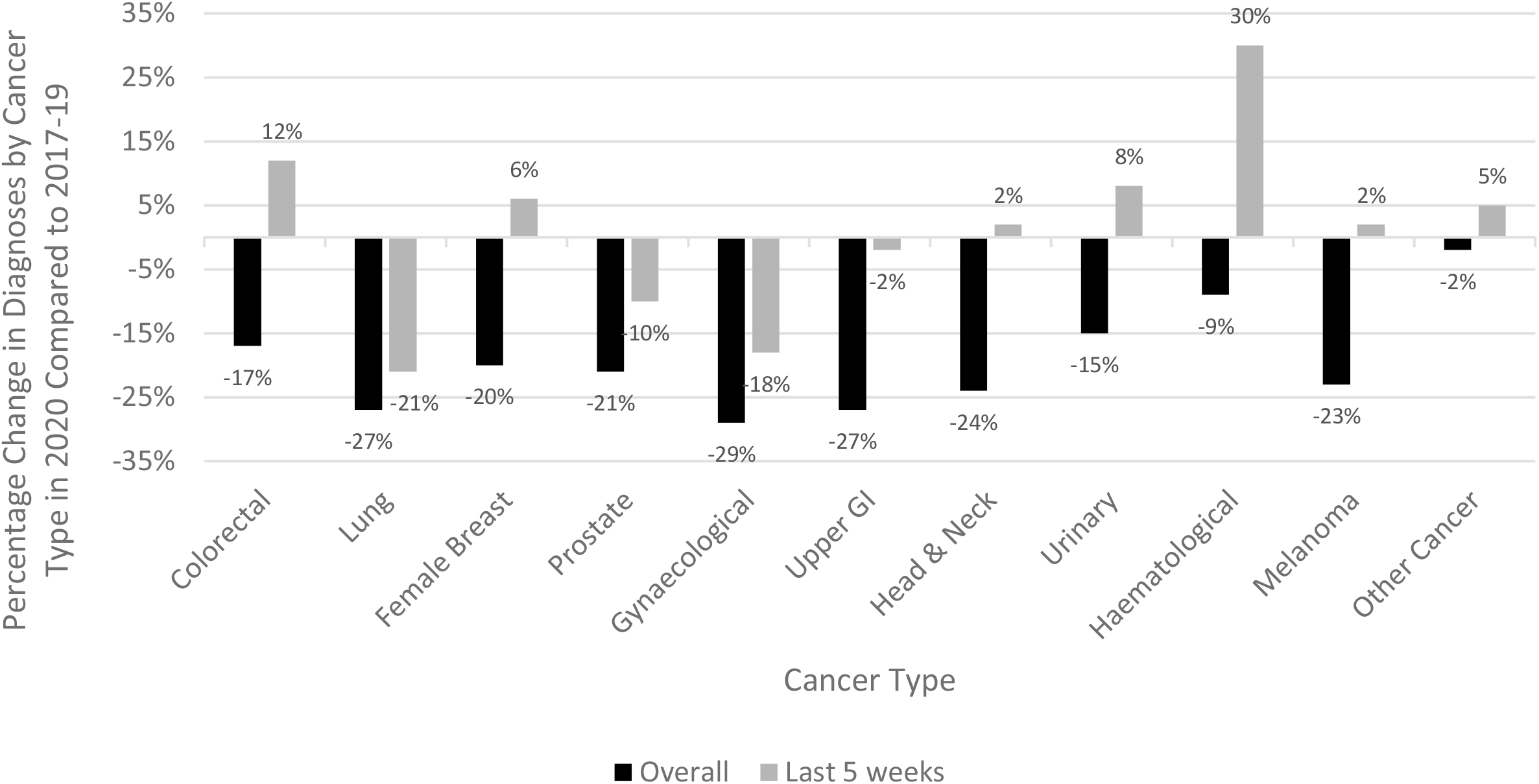
Percentage change in diagnoses by cancer type, overall from 1st March to 12th September 2020, and from the most recent 5 week period studied from 9th August to 12th September 2020, compared to the same time periods in 2017-19.

### Analysis by screen-detected colorectal and breast cancers

Trends for colorectal cancer and breast cancer for the general population and for the screening age population (60-74 years for colorectal cancer and 50-70 years for breast cancer) are shown in Figure 3. The number of colorectal cancer diagnoses was reduced for all ages and for the screening age population during the overall six month period studied compared to previous years. From 9^th^ August to 12^th^ September 2020 the total number of colorectal cancer diagnoses increased by 12%, compared to previous years, indicating some recovery for the population as a whole. However, in the screening age population, the number of diagnoses remained significantly reduced, by 18% lower than previous years. The number of breast cancer diagnoses was reduced for both the general and screening age population during the overall six month period studied in 2020 compared to previous years. In the more recent five week period studied however, recovery was evident in both groups, with an increase of 6% overall and 14% in the screening age group, in the number of breast cancer cases compared to previous years.

**Figure 3.**
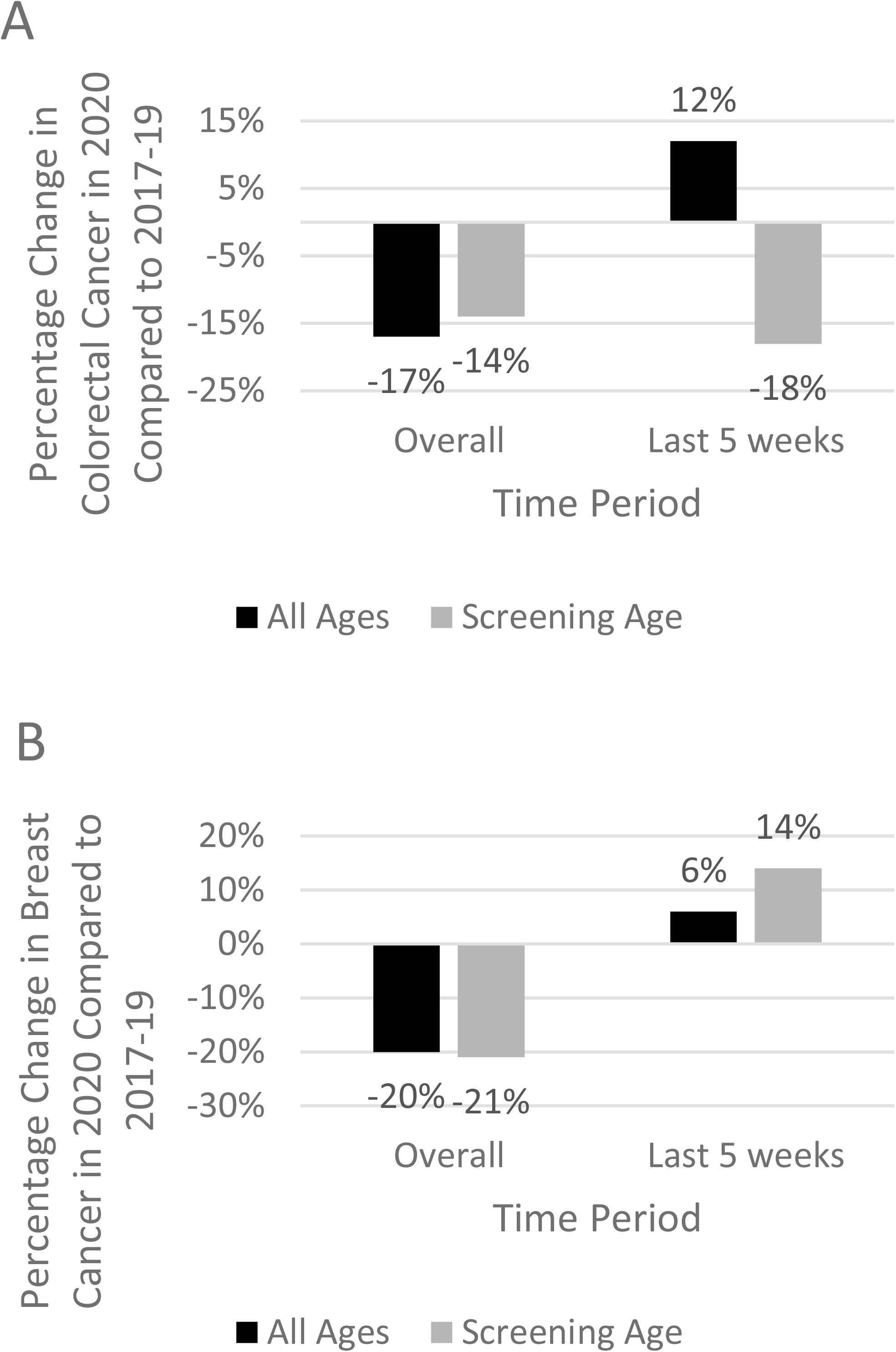
Percentage change in colorectal and breast cancer diagnoses, overall from 1st March to 12th September 2020 and the most recent time period studied from 9th August to 12th September 2020, for all ages and for the screening age population, compared to the same time periods in 2017-19.

### Analysis by age and sex

Both sexes were relatively equally affected overall, with a 22% and 23% reduction in cancer diagnoses in males and females during the overall pandemic time period (Figure 4). However, when studying the most recent five week time period from 9^th^ August to 12^th^ September 2020, males lagged somewhat behind females, with observed 8% and 2% lower numbers of pathological cancer diagnoses, respectively.

**Figure 4.**
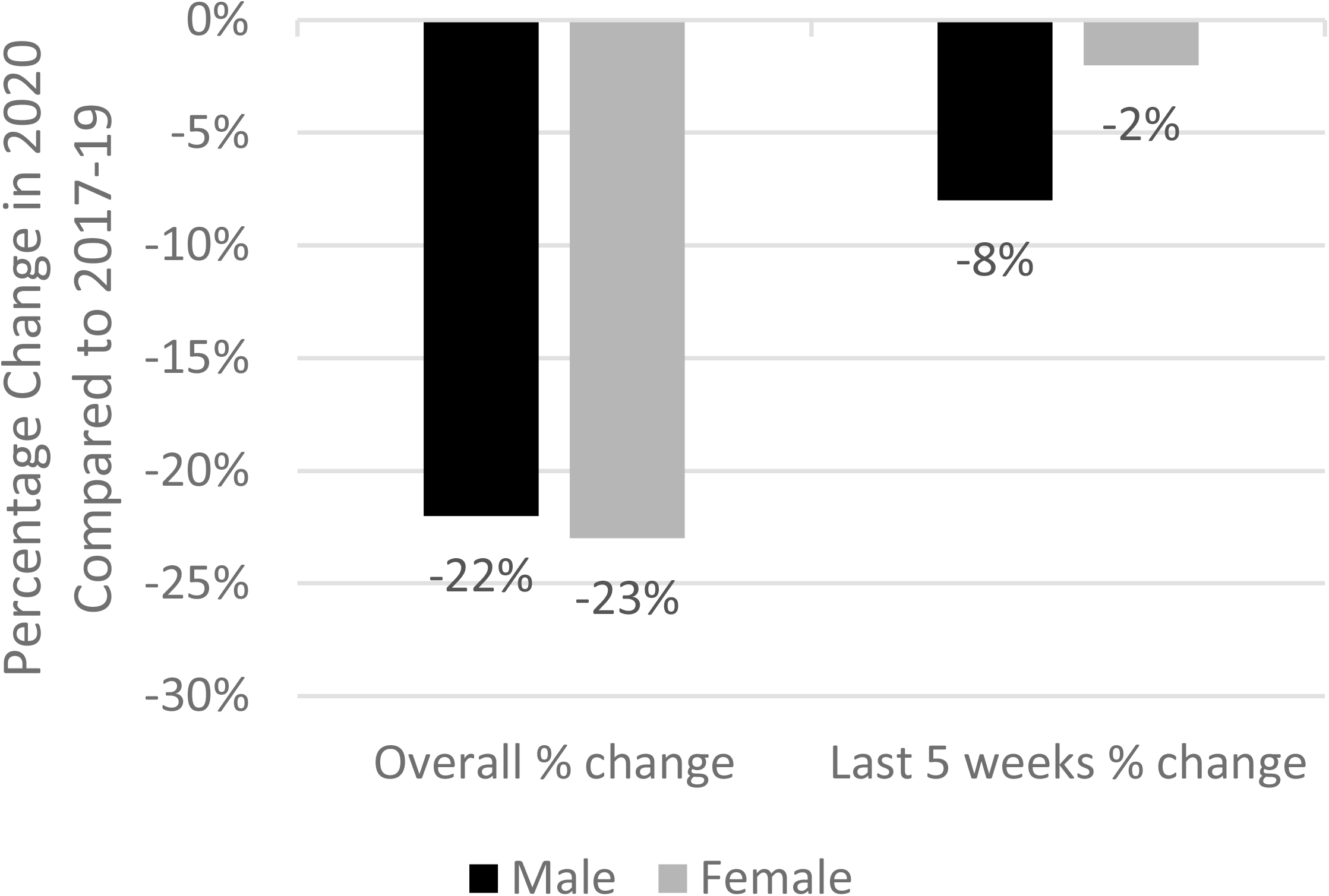
Percentage change in cancer diagnoses by gender, overall from 1st March to 12th September 2020 and the most recent time period studied from 9th August to 12th September 2020, compared to the same time periods in 2017-19.

The number of cancer diagnoses across age categories recorded from 1^st^ March to 12^th^ September 2020 is shown in Figure 5A. The largest proportional decrease in pathology samples indicating cancer was in the 50-59 years old age group, in which cancer cases reduced by 27.4% in 2020 compared to previous years. The number of cancer diagnoses per month by age category are shown in Figures 5B-F, both during the first wave of the COVID-19 pandemic of 2020 and the average number per month from 2017-2019 for comparison. Some recovery is shown over the summer months, although not to pre-pandemic levels in any age group. While the relative decrease in cancer diagnoses was greatest for the 50-59 years age group, the absolute decrease in the number of cancer cases was greatest in the 70-79 years age group (275 cases) followed by the 60-69 years age group (229 cases) and the 50-59 years group (217 cases).

**Figure 5.**
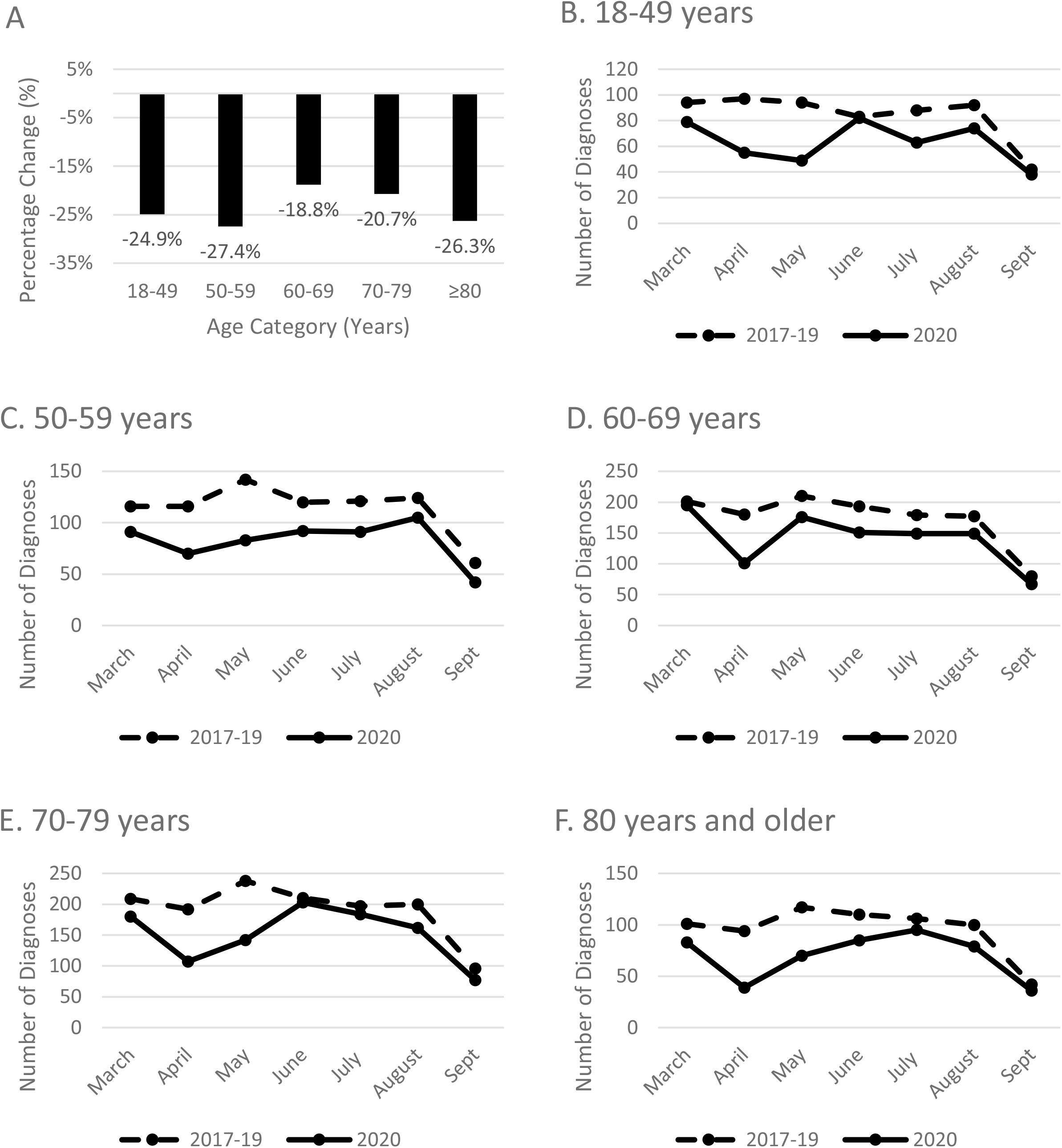
A Percentage change in cancer diagnoses by age category from 1st March to 12th September 2020 compared to the same time period in 2017-19. Note the apparent decline in September reflects that data was not available for the whole month. B-F The number of cancer diagnoses per month by age category in 2020 and 2017-19. B 18-49 years. C 50-59 years. D 60-69 years. E 70-79 years. F 80 years and older

## Discussion

Our data demonstrate the considerable effect that the COVID-19 pandemic has had on reducing the number of people being pathologically diagnosed with cancer in Northern Ireland. We were able to quantify this as a 23% drop in expected pathological cancer diagnoses in Northern Ireland during the first six months of the COVID-19 pandemic taking hold in the UK, translating to 1,130 ‘missed’ cancer patients over this time period.

Likely causation is complex, with factors involving patients, primary care and secondary care all contributing. Some cases are likely due to failure by the patient to report concerning symptoms for investigation. Failure to engage with the healthcare system may reflect patient fear regarding contracting COVID-19, or an effort on their part to “protect the NHS”, given national guidance to this effect, or limited ability to use telephone consultations.^(3, 14)^ Primary care has seen a shift away from face-to-face consultations towards telephone consultations,^(15)^ which may have led to missed detection of concerning symptoms or signs, or to a delay in appropriate diagnostic investigations such as radiology or endoscopy. In secondary care the capacity for diagnostic investigations has been limited by COVID-19, both in terms of resources and staffing levels, as well as the need for infection control precautions.^(16)^ Staff absence due to illness or self-isolation is likely to also be a contributing factor in both primary and secondary care, reducing the number of patient assessments able to be carried out. It should be noted that a proportion of these ‘missed’ patients may have been diagnosed clinically instead of pathologically (for example, as a result of an emergency admission to hospital) and, if their subsequent treatment did not generate a diagnostic pathology specimen, such cases will be excluded from the data presented. However, these cases are likely to be a small number of the total.

Some recovery in cancer diagnoses has occurred during the 2020 summer months, but this has revealed important inequalities in recovery across cancer sites and population groups. In the more recent time period studied, some recovery in pathological cancer diagnoses was observed across most cancer sites. However, lung, gynaecological and prostate cancers continue to lag behind the expected number of pathological diagnoses in 2020 compared to the preceding three years. The reasons behind this are unclear, and are likely multifactorial. Most concerningly, reduced lung cancer diagnoses may reflect an overlap and confusion with COVID-19 symptoms, and ‘stay at home’ advice for individuals with persistent coughs, rather than referral for radiological investigation. In addition, a nationwide survey of endoscopy units in Germany in April 2020 revealed that two thirds of units had cancelled at least 20% of scheduled bronchoscopy procedures.^(17)^ Bronchoscopy is an aerosol-generating procedure which carries a risk of transmitting COVID-19 infection. The British Thoracic Society has made recommendations to mitigate this, which includes triage guidelines for patients, comprising symptom enquiry and COVID-19 testing, along with personal protective equipment for healthcare professionals,^(18)^ both of which could contribute to lower numbers of bronchoscopies being done.

Little data exist on the impact of the COVID-19 pandemic on specific cancer sites to date. A hospital-based case series from South Korea reported that the number of lung cancer cases diagnosed from February to June 2020 did not differ significantly from the previous three years.^(9)^ However, this study did also report that the number of stage III-IV non-small-cell lung cancer cases was significantly higher in 2020 compared to previous years (74.7% compared to 57.9-66.7% in 2017-19).^(9)^ Unfortunately we do not have staging information for the cancer data presented in this article as yet, but a shift in distribution to later stage at presentation is likely to be seen for cancer patients in the coming months and years. There is limited generalisability between the UK and South Korea in terms of experience during the COVID-19 pandemic. Nevertheless, it is concerning and predicted that these ‘missed’ cancer patients are likely to present with a more advanced cancer, and therefore have poorer outcomes, at some point in the future.

The increased number of breast cancer diagnoses from 9^th^ August to 12^th^ September 2020 for the whole population, and in those of screening age, suggests the successful resumption of cancer screening is contributing to the observed recovery in numbers of cancer diagnoses. In contrast to investigation of other cancers, such as lung cancer, diagnosis of breast cancer does not require an aerosol-generating procedure and personal protective equipment, therefore capacity for breast cancer investigations is likely to be nearer normal pre-pandemic levels. These results are in keeping with trends in the Netherlands, which showed a steep decline in breast cancer cases during the initial stages of the pandemic, with the screening age population more affected than women of non-screening age.^(8)^ However, in the Netherlands the non-screening age group recovered more quickly to expected levels, whereas our results suggest a more rapid recovery in the screening age group.

In contrast to breast cancer screening which resumed in mid-July 2020, colorectal cancer screening invitations did not resume in Northern Ireland until mid-August 2020. Furthermore, the colorectal cancer screening pathway involves the secondary step of colonoscopy prior to cancer diagnosis and cessation of non-urgent endoscopy at the height of the pandemic generated a significant and ongoing backlog of individuals awaiting screening colonoscopy. Therefore recovery of diagnoses in screen-detected colorectal cancers is not yet evident within the timeframe of this study.

Pathological cancer diagnoses in males have shown less recovery than females in recent weeks. This finding is in keeping with prostate cancer diagnoses lagging behind some other cancer sites. While all age groups have been affected, the reduction in cancer cases varied from -18.8% to -27.4%, with the worst affected group being the 50-59 year age group. Two UK-based studies of adults aged 50 years and older found that women were more likely than men to consult their GP regarding symptoms possibly indicative of cancer,^(19)^ and that people of an older age (aged 60-69 compared to the youngest age group) were more likely to have sought help for cancer ‘alarm’ symptoms.^(20)^ Our findings are in keeping with this. However, how interactions with healthcare providers during the COVID-19 pandemic vary across gender and age categories remains mostly unknown. The reasons for observed disparities in gender and age require evaluation in other population settings to understand if these trends are occurring elsewhere. Targeted public health campaigns may be required to reduce such inequalities as the second wave of the COVID-19 pandemic continues to grow across the UK and internationally.

Health professionals recognise the need to rebuild and protect cancer services during the ongoing COVID-19 pandemic, particularly in the areas of diagnostic procedures and surgery.^(21)^ Communicating key messages to the public around awareness of cancer symptoms and uptake of screening invitations is of utmost importance, along with emphasis on seeking medical attention if required. An example of this is the NHS “Open for business” campaign run by Public Health England earlier this year.^(22)^ Social media has the potential to be effective in cancer prevention^(23)^ and could be employed for public health messaging, particularly for younger adults. Communication directed at older age groups could comprise more traditional media channels, but given the unique challenges regarding health communication globally in 2020,^(24)^ the optimal mode of campaigning remains to be determined.

A key strength of this study is its population coverage. The NICR was able to mobilise and alter their working practices in the initial stages of the pandemic, to retrieve rapid data on pathological cancer diagnoses in order to assess the impact of COVID-19 across the region. Limitations include basing our data on pathology diagnoses rather than following usual cancer registration methods, and thus have not been as rigorously validated. Further pathology samples taken during January to September 2020 may not yet have been recorded if a result was not available by the end of September 2020. Therefore, the presented figures may represent an underestimation, particularly for later weeks. However, the data for 2017-2019 was also based on pathological samples indicating cancer, in order to provide consistency and to attempt to control for solely clinical diagnoses. The interpretation of the trends observed in the most recent 5 week time period studied, the 9^th^ August to the 12^th^ September, should be interpreted with caution given the instability of health services and rapid changes in restrictions seen during the pandemic so far, as numbers may change on a monthly basis. Prior to the submission of this article, we obtained a brief update for the 5 week period from 6^th^ September to 10^th^ October 2020, which showed that while lung and prostate cancer diagnoses remained 30% and 19% below the level of previous years, diagnoses of gynaecological cancers appeared to have recovered to near expected levels. In addition, some cancer sites such as melanoma, head & neck and upper GI cancers have seen the numbers of diagnoses reduce by 37%, 27% and 23% respectively, despite showing recovery in the previous 5 week period studied. This illustrates the unpredictable nature of cancer diagnostics during an international pandemic, the need for ongoing data collection, and the realisation that the impact of COVID-19 won’t be fully understood until several more months, if not years, have passed. Nevertheless, the need for timely understanding of the impact of the COVID-19 pandemic on cancer diagnoses has required such approaches to be undertaken.

In summary, this population-based study has quantified the impact of the COVID-19 pandemic on reducing by 23% the number of cancer diagnoses during the first six months of the pandemic in a UK region. There are emerging inequalities by cancer site and population subgroups in cancer diagnoses which require further investigation to optimise recovery pathways. Additional variables, such as socioeconomic status, may also be contributing to the observed inequalities, and could be included in future research. As the COVID-19 pandemic continues, measures to mitigate the impact on cancer diagnoses must be taken in the areas of public education and awareness campaigns, rapid access to healthcare professionals, referral pathways and diagnostic capacity. It is expected that the observed reduction in cancer diagnoses, and particularly screen-detected cancers, will translate into diagnosis at more advanced stages of disease for those patients in whom investigations were delayed, and therefore poorer survival outcomes. Longer-term data are needed, including information on cancer site, stage, treatment and outcomes, to fully assess the impact of COVID-19 on cancer care, and to assist with ongoing service recovery planning.

## Data Availability

Monthly reports of data regarding cancer diagnoses in Northern Ireland, on which this study was based, are available on the Northern Ireland Cancer Registry website.

## Additional Information

### Acknowledgements

This research has been conducted using data from the Northern Ireland Cancer Registry (NICR) which is funded by the Public Health Agency, Northern Ireland. However, the interpretation and conclusions of the data are the sole responsibility of the author(s). The author(s) acknowledge the contribution of the NICR staff in the production of the NICR data. Like all Cancer Registries our work uses data provided by patients and collected by the Health service as part of their care and support. Richard Turkington is supported by Cancer Research UK (C50880/A29831), Cancer Focus NI and OGCancer NI, Helen Coleman is supported by Cancer Research UK (C37703/A25820). Ashleigh Hamilton is supported by the HSC R&D Division, Public Health Agency, Northern Ireland (EAT/5494/18).

## Authors’ Contributions

Study conception and design: HGC, ACH

Data acquisition: DWD, CF, DF

Data analysis and interpretation: HGC, ACH, MBL, RCT, DWD, ATG

Drafting manuscript: HGC, ACH, MBL, RCT, ATG, DWD, CEO’N

Manuscript revision and final approval: All

## Ethics Approval and Consent to Participate

Ethical approval for the NICR databases (including the waiving of requirement for individual patient consent) and data analysis has been granted by the Office for Research Ethics Committees of Northern Ireland (ORECNI reference 15/NI/0203), recently renewed in October 2020 (REC REF 20-NI-0132).

## Consent for Publication

Not applicable – no identifiable individual data in manuscript.

## Data Availability

Monthly reports of data regarding cancer diagnoses in Northern Ireland, on which this study was based, are available on the Northern Ireland Cancer Registry website.^(13)^

